# Multidrug resistance bacteria causing community acquired urinary tract infections among adult outpatients attending lower-level health facilities in Mwanza, Tanzania

**DOI:** 10.64898/2025.11.30.25341325

**Authors:** Eunice G. Emmanuel, Ashura Khamis, Elikana Michael, Dorina Muhizi, James Thomas, Bernard Okamo, Farida I. Mkassy, Eveline T. Konje, Vitus Silago, Martha F. Mushi, Stephen E. Mshana

**Author notes:** Correspondence: Martha F. Mushi.

## Abstract

**Background:** Urinary tract infections (UTIs) remain a common clinical condition requiring antibiotic prescription among adult outpatients in low- and middle-income countries (LMICs). Data regarding the prevalence, patterns of bacteria and their antibacterial susceptibility profile for community acquired UTI are limited.

**Methods:** A hospital-based cross-sectional study was conducted from May to August 2024 at Igoma and Buzuruga Health Centres in Mwanza. Symptomatic adults’ patients, without recent hospitalization (within past 30 days) or urinary catheterization were enrolled. Clinical and demographic data were collected followed by standard mid-stream urine culture and disc diffusion susceptibility testing. Descriptive data analysis was performed using STATA version 15.

**Results:** A total of 1,005 adult patients with the median age of 32 [IQR: 23–49] years were recruited, of whom 727 (72.3%) were female. The majority (64.3%) reported a previous history of UTI (within six months), and the median symptom duration before presentation was 8 [5–15] days. Microbiological confirmation of UTI was found in 221 patients (22.0%, 95% CI 19.5%-24.7%). The most frequently isolated uropathogens were *Escherichia coli* (32.9%) and *Staphylococcus aureus* (24.7%). *Escherichia coli* isolates were ≥50% resistant to ciprofloxacin, amoxicillin-clavulanic acid, trimethoprim-sulfamethoxazole, tetracycline, and ampicillin. About 20.3% of *E. coli* isolates showed positive extended-spectrum beta-lactamase (ESBL) phenotypes whereas 64.6% of *S. aureus* were resistant to cefoxitin, hence methicillin-resistant *S. aureus* (MRSA) strains.

**Conclusion and recommendation:** This study revealed a significant burden of community-acquired urinary tract infections (CA-UTIs) among adult outpatients, with an overall prevalence of 22%. *Escherichia coli* and *Staphylococcus aureus* were the most frequently isolated pathogens, and alarmingly high levels of antimicrobial resistance, particularly multidrug resistance in *E. coli*. Improved diagnostic capacity and strengthened antibiotic stewardship are urgently needed to guide effective management of community-acquired UTIs to control the AMR development.

**Highlight:**

- A total of 1,005 symptomatic adult patients at primary healthcare facilities were assessed for community-acquired urinary tract infections (CA-UTIs).
- CA-UTIs were identified in 22% of cases, with *Escherichia coli* and *Staphylococcus aureus* as the most common pathogens.
- *E. coli* demonstrated a notably high rate of multidrug resistance, with 82.8% of isolates resistant to multiple antibiotic classes.
- Over 70% of Gram-negative bacterial isolates were resistant to at least three classes of antibiotics, presenting major treatment difficulties.
- These results highlight the urgent need for ongoing surveillance and updated empirical treatment policies for UTIs in outpatient clinics, especially in resource-limited settings.

## Background

Urinary tract infections (UTIs) are prevalent health concern among patients visiting outpatient clinics in low- and middle-income countries (LMICs). In these regions, UTIs significantly contribute to morbidity, elevated treatment expenses, and mortality, especially among those seeking care at lower-tier health facilities [1, 2]. Limited diagnostic resources coupled with insufficient microbiological evidence often compromise effective patient management. UTI is the most common reason for AB prescription and the most frequent reason for hospital attendance among children and adults in Tanzania surpassed only by malaria [3, 4]. In Tanzania, the prevalence of UTI is reported to be different among populations ranging from 11-23% [5–7]. A previous study conducted in lower health facilities in Tanzania among patients of all age groups, reported the culture proven prevalence of 27.4% [8, 9].

Gram negative bacteria in the order Enterobacterales such as *E. coli* and *Klebsiella pneumoniae* are the most common uropathogens associated with both community and health care settings UTIs [1, 9]. These isolates have been found to produce extended spectrum beta-lactamase (ESBL).Among Gram positive bacteria, *Staphylococcus aureus* has been reported as the leading pathogen [10]. In LMICs studies have documented enzyme production like extended spectrum beta-lactamase (ESBL) as the main mechanism of resistance among Gram negative bacteria causing UTIs [11, 12]. Previous studies conducted in Tanzania reported the prevalence of multi drug resistant (MDR) bacteria causing UTIs to range from 22.4%-60.3% among Gram positive bacteria and 45.4%-52.2% among Gram negative bacteria [4, 9, 13, 14].

Several factors increase the risk of community-acquired urinary tract infections (CA-UTIs). Demographic and physiological factors are significant, with women being more vulnerable due to their shorter urethra and its proximity to the anus. Older adults are at greater risk due to weakened immune defences and underlying health conditions, while pregnancy increases susceptibility because of hormonal changes and pressure on the bladder[15]. Behavioural and lifestyle factors, such as poor hygiene, frequent sexual activity, and the use of spermicides or diaphragms, can disrupt the natural bacterial balance in the urinary tract, further increasing the likelihood of infection.

Additionally, certain medical conditions and structural abnormalities heighten the risk. Individuals with diabetes mellitus are more prone to UTIs due to compromised immune function and elevated glucose levels that encourage bacterial growth. Conditions such as kidney stones, congenital abnormalities, or an enlarged prostate can obstruct urine flow, fostering an environment for bacterial proliferation. People with history of recurrent UTIs are also at higher risk for future infections. Medications and medical interventions are other contributing factors, as the use of antibiotics can disturb the balance of normal flora, and temporary catheterization, even at home, can increase the likelihood of bacterial colonization[4].

This study was conducted to document the prevalence, factors associated and profile of bacteria causing CA-UTIs. The antimicrobial susceptibility pattern of these uropathogens are well depicted.

## Methodology

### Study design and settings

This was a hospital-based cross-sectional study conducted from 15^th^ May to 30^th^ August 2024, at Igoma and Buzuruga Health Centre in Mwanza, Tanzania. Buzuruga Health Centre serves up to 2,266 patients per month while Igoma Health Centre attend approximately 1,500 outpatients monthly.

### Sample size and study population

The study involved symptomatic adult patients presenting with signs and symptoms of UTIs. To be enrolled in the study, the patient was supposed to present with at least two of the following UTI symptoms; dysuria, frequency, urgency, nocturia, suprapubic pain, haematuria, and fever. The minimum sample size was 306 symptomatic adults obtained by Kish-Leslie formula [16] using a 24.7% prevalence of CA-UTIs previously reported by Silago et *al*.[9]. Pregnant women, patients with history of admission within 30 days on the day of sample collection and those with an indwelling urinary catheter at the time of study were excluded.

### Data and sample collections

Demographic and clinical data were collected using epicollect5 a mobile data-gathering platform developed by the CGPS Team of Oxford BDI and publicly available at https://five.epicollect.net. About 3-5 ml of clean catch midstream urine was collected by the patients after receiving instructions from research assistant, using a clean, wide-mouthed, labelled container. The urine sample was transported to microbiology research laboratory of CUHAS for processing within 2 hours of collection. For women, urine samples were tested for HCG to exclude pregnancy. Urine was divided to two portions for microscopy/dipstick analysis and culture. All laboratory data were documented on the laboratory logbook.

### Urine Culture

Urine samples were inoculated onto agar media (Blood Agar, MacConkey Agar and CLED) and incubated at 37°c for 18-24 hours. Growth of ≥1000 colonies from 10µl inoculating loop for both men and women was considered as significant bacteriuria and thus UTI [17]. In-house-prepared conventional biochemical identification tests were used for the preliminary identification of bacteria isolates to their possible species levels as previously described [18]. Gram-positive and Gram-negative bacteria were identified using morphological and biochemical tests as previously described [19]. Unidentified isolates were further analysed using Matrix-Assisted Laser Desorption Ionization–Time of Flight (MALDI-TOF) mass spectrometry at the National Public Health Laboratory (NPHL). Antimicrobial susceptibility testing was performed as per CLSI guidelines [20].

### Quality Control

Quality assurance measures encompassed monitoring reagent storage, equipment functionality, media sterility, and preparation of standardized bacterial suspensions. All stains and reagents were systematically labelled, dated, and stored under appropriate conditions. Refrigerator and incubator temperatures were recorded and tracked daily to ensure compliance with operational standards. Culture media were prepared in strict accordance with manufacturer instructions and evaluated for both performance and sterility. To maintain consistent inoculum density for susceptibility testing, a 0.5 McFarland standard was utilized; standard reference strains like *S. aureus* (ATCC 25923), *MRSA* (ATCC 29213), *E. coli* (ATCC 25922), ESBL-producing *Klebsiella pneumoniae* (ATCC 700603), and *Pseudomonas aeruginosa* (ATCC 27853) were employed as controls. Data cleaning and coding were conducted using STATA version 15, with records containing missing values excluded from analysis.

### Data Analysis

Data were analysed using STATA software version 15 in line with the study’s objectives. Continuous (age and duration of symptoms) and categorical variables (gender, education, marital status, clinical presentation, isolates and antimicrobial susceptibility testing profile) were summarized using medians and proportions, respectively.

### Ethical Considerations

Ethical clearance was obtained from the Joint CUHAS/BMC Research Ethics and Review Committee (CREC) with certificate number CREC No 763/2024. All participants were requested to consent by signing written informed consent form. Privacy and confidentiality were maintained throughout the study. The urine culture results were timely shared to the manage clinicians to guide the management.

## Results

The study recruited a total of 1005 adult, patients with a clinical diagnosis of UTI. In general, majority of recruited patients were non-pregnant women 727(72.3%), about two third were married 610 (60.7%) and their median age was 32[23–49] years, as detailed in table1.

In general, about two thirds of the patients had previous history of UTI diagnosis (within past six months) 646(64.3%), and the median duration of the current symptoms before reporting to the health facility was 8[5–15] days. Patients recruited from Igoma health centre reported late to the health facility than those from Buzuruga health centre (14[7–28] vs. 7[3–14] days), P=0.001.

Though more than half, 588(58.5%), reported to have fever (>=38.5 °C) only few reported to have classical symptoms of UTI like dysuria 254(25.3%), urgency 70(6.9%) cloudy urine 89(8.9%) and smelling urine 54(5.4%). Prior antibiotic use (within a week) for treatment of the current symptoms was reported by 12.8% (129) patients, as indicated in table 2.

**Table 1:** Social demographic characteristics of the studied patients.

| Variable |  | Buzuruga HC (566)<br>n(%) | Igoma HC (439)<br>n(%) | Total (1005) |
| --- | --- | --- | --- | --- |
| Gender | Male | 138(24.4) | 140(31.9) | 278(27.7) |
|  | Female | 428(75.6) | 299(68.1) | 727(72.3) |
| Age | Median [IQR] | 32[23-50] | 30[22-49] | 32[23-49] |
| Education | No formal education | 37(6.5) | 65(14.8) | 102(10.2) |
|  | Primary | 243(42.9) | 216(49.2) | 459(45.7) |
|  | Secondary | 220(38.9) | 123(28.0) | 343(34.1) |
|  | University | 66(11.7) | 35(7.9) | 101(10.1) |
| Marital status | Married | 367(64.8) | 243(55.4) | 610(60.7) |
|  | Single | 199(35.2) | 196(44.6) | 395(39.3) |

**Table 2:**
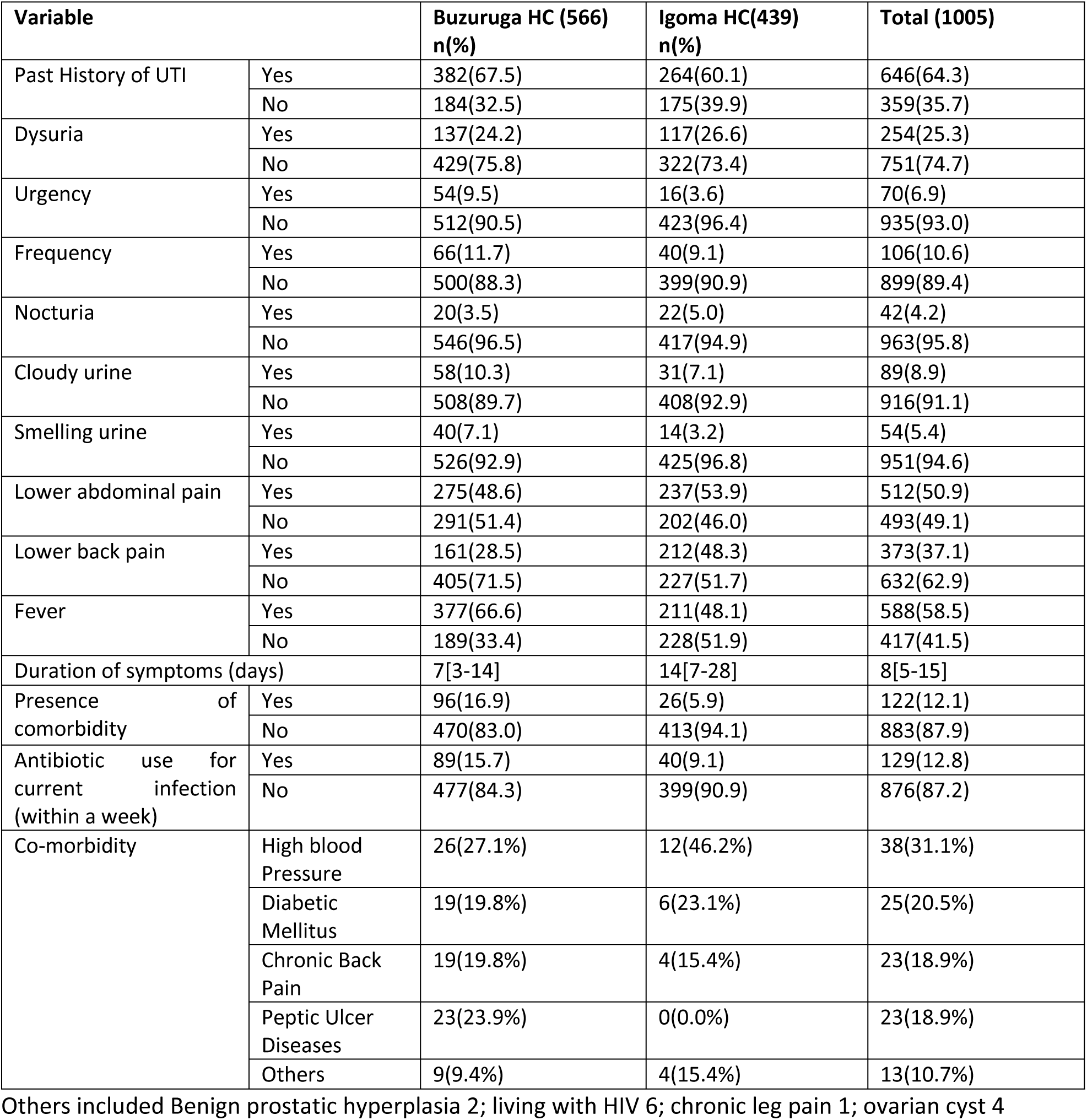
Clinical characteristics of the recruited patients.

| Variable |  | Buzuruga HC (566)<br>n(%) | Igoma HC(439)<br>n(%) | Total (1005) |
| --- | --- | --- | --- | --- |
| Past History of UTI | Yes | 382(67.5) | 264(60.1) | 646(64.3) |
|  | No | 184(32.5) | 175(39.9) | 359(35.7) |
| Dysuria | Yes | 137(24.2) | 117(26.6) | 254(25.3) |
|  | No | 429(75.8) | 322(73.4) | 751(74.7) |
| Urgency | Yes | 54(9.5) | 16(3.6) | 70(6.9) |
|  | No | 512(90.5) | 423(96.4) | 935(93.0) |
| Frequency | Yes | 66(11.7) | 40(9.1) | 106(10.6) |
|  | No | 500(88.3) | 399(90.9) | 899(89.4) |
| Nocturia | Yes | 20(3.5) | 22(5.0) | 42(4.2) |
|  | No | 546(96.5) | 417(94.9) | 963(95.8) |
| Cloudy urine | Yes | 58(10.3) | 31(7.1) | 89(8.9) |
|  | No | 508(89.7) | 408(92.9) | 916(91.1) |
| Smelling urine | Yes | 40(7.1) | 14(3.2) | 54(5.4) |
|  | No | 526(92.9) | 425(96.8) | 951(94.6) |
| Lower abdominal pain | Yes | 275(48.6) | 237(53.9) | 512(50.9) |
|  | No | 291(51.4) | 202(46.0) | 493(49.1) |
| Lower back pain | Yes | 161(28.5) | 212(48.3) | 373(37.1) |
|  | No | 405(71.5) | 227(51.7) | 632(62.9) |
| Fever | Yes | 377(66.6) | 211(48.1) | 588(58.5) |
|  | No | 189(33.4) | 228(51.9) | 417(41.5) |
| Duration of symptoms (days) |  | 7[3-14] | 14[7-28] | 8[5-15] |
| Presence of comorbidity | Yes | 96(16.9) | 26(5.9) | 122(12.1) |
|  | No | 470(83.0) | 413(94.1) | 883(87.9) |
| Antibiotic use for current infection (within a week) | Yes | 89(15.7) | 40(9.1) | 129(12.8) |
|  | No | 477(84.3) | 399(90.9) | 876(87.2) |
| Co-morbidity | High blood Pressure | 26(27.1%) | 12(46.2%) | 38(31.1%) |
|  | Diabetic Mellitus | 19(19.8%) | 6(23.1%) | 25(20.5%) |
|  | Chronic Back Pain | 19(19.8%) | 4(15.4%) | 23(18.9%) |
|  | Peptic Ulcer Diseases | 23(23.9%) | 0(0.0%) | 23(18.9%) |
|  | Others | 9(9.4%) | 4(15.4%) | 13(10.7%) |

Others included Benign prostatic hyperplasia 2; living with HIV 6; chronic leg pain 1; ovarian cyst 4

Among the recruited patients with other comorbidities 122, high blood pressure was the commonest, 38 (31.1%). A total of seven patients had more than one chronic condition (comorbidity) such as diabetic mellitus and hypertension (six patients), and one patient had both having hypertension and peptic ulcer diseases making total count of 122.

### Prevalence of community acquired UTI

Of the 1005 recruited patients with clinical presentation of urinary tract infections, 221(22%), 95% CI 19.4%-24.6% were microbiologically confirmed. Igoma health centre had 22.8%, (n=100/439), 95% CI 18.9%-27% patients with microbiologically confirmed UTI while Buzuruga health centre had 21.4%, (n=121/566), 95% CI 18%-25% patients.

### Proportions of bacterial species causing CA-UTIs

A total of 194 bacterial isolates were identified from significant positive urine samples, with the majority being Gram-negative bacteria (59.8%; n = 116). Overall, the most frequently detected bacterial species were *E. coli* (32.9%; n = 64) and *S. aureus* (24.7%; n = 48). A similar pattern of predominant bacteria pathogens was observed at both Buzuruga Health Centre and Igoma Health Centre, however, the third and fourth most common isolates differed slightly between the two sites. At Buzuruga HC, *Enterococcus* spp. (10.5%; n = 11) and *Acinetobacter* spp. (9.5%; n = 10) were the commonest third and fourth pathogen, respectively while at Igoma *Acinetobacter* spp. (10.1%; n = 9) and *K. pneumoniae* complex (8.9%; n = 8) were common. Notably, all *C. freundii* strains were exclusively isolated at Buzuruga HC (Table 3//Figure 2).

**Figure 2:**
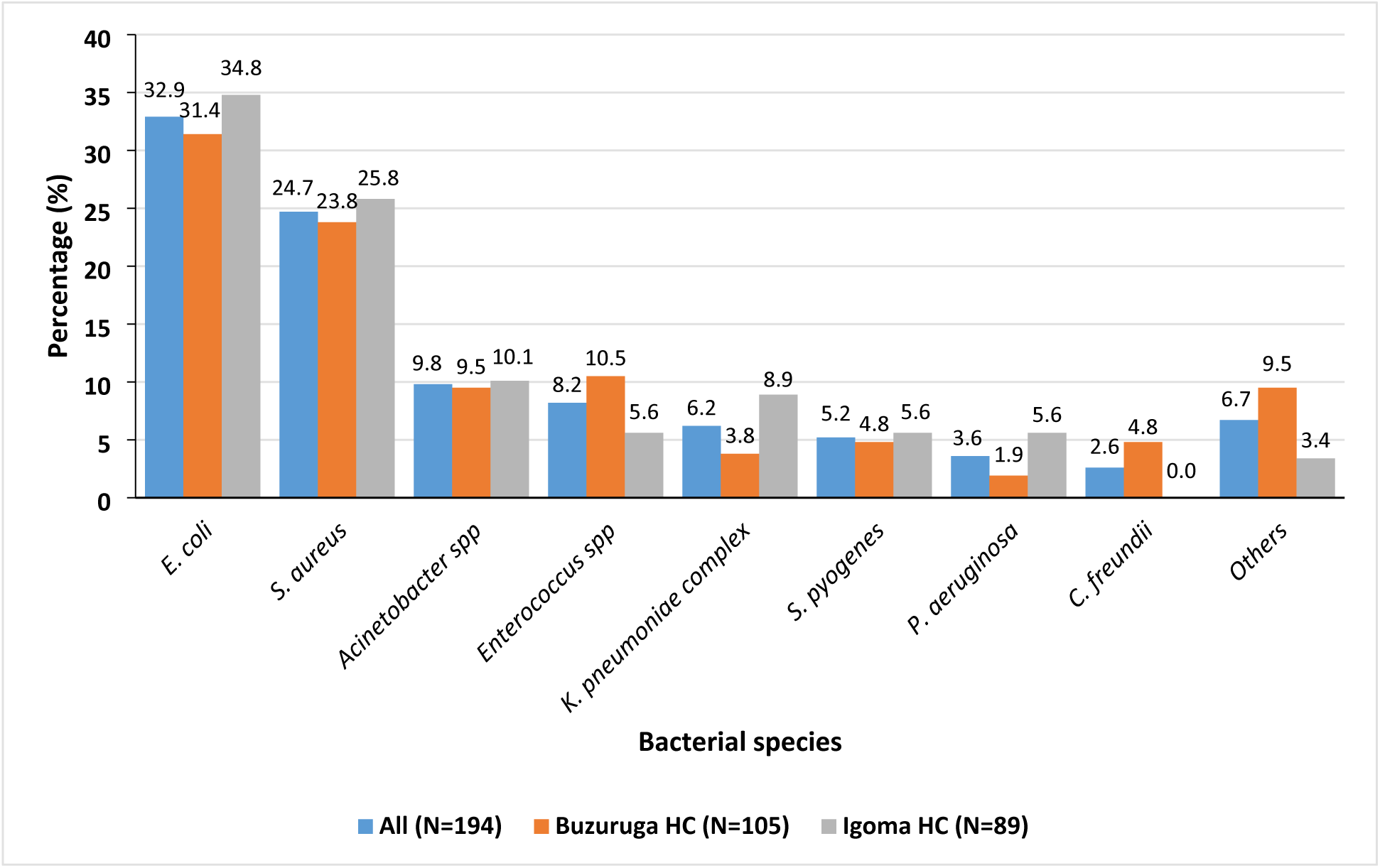
The proportions of bacterial species causing community-acquired urinary tract infections at Buzuruga and Igoma Health Centers in Mwanza, Tanzania. ***\**All (n=13):** *Morganella morganii* (n=3), *Enterobacter cloacae* complex (n=2), *Citrobacter koseri* (n=1), *Klebsiella oxytoca* (n=1), *Pseudomonas fluorescens* (n=1), *Proteus mirabilis* (n=1), *Pseudoglutamicibacter cumminsii* (n=1), *Streptococcus agalactiae* (n=1), *Staphylococcus epidermidis* (n=1), and *Staphylococcus haemolyticus* (n=1). **Buzuruga HC (n=10):** *Morganella morganii* (n=3), *Enterobacter cloacae* complex (n=1), *Citrobacter koseri* (n=1), *Klebsiella oxytoca* (n=1), *Pseudomonas fluorescens* (n=1), *Pseudoglutamicibacter cumminsii* (n=1), *Staphylococcus epidermidis* (n=1), and *Staphylococcus haemolyticus* (n=1). **Igoma HC (n=3):** *Enterobacter cloacae* complex (n=1), *Proteus mirabilis* (n=1), and *Streptococcus agalactiae* (n=1).

**Table 3:** The proportions of bacterial species causing community-acquired urinary tract infections at Buzuruga and Igoma Health Centers in Mwanza, Tanzania.

| Isolate | All (n=194) | Buzuruga HC (N=105) | Igoma HC (N=89) |
| --- | --- | --- | --- |
|  | n(%) | n(%) | n(%) |
| *E. coli* | 64 (32.9) | 33 (31.4) | 31 (34.8) |
| *S. aureus* | 48 (24.7) | 25 (23.8) | 23 (25.8) |
| *Acinetobacter* spp. | 19 (9.8) | 10 (9.5) | 9 (10.1) |
| *Enterococcus* spp. | 16 (8.2) | 11 (10.5) | 5 (5.6) |
| *K. pneumoniae* complex | 12 (6.2) | 4 (3.8) | 8 (8.9) |
| *S. pyogenes* | 10 (5.2) | 5 (4.8) | 5 (5.6) |
| *P. aeruginosa* | 7 (3.6) | 2 (1.9) | 5 (5.6) |
| *C. freundii* | 5 (2.6) | 5 (4.8) | 0 (0.0) |
| Others* | 13 (6.7) | 10 (9.5) | 3 (3.4) |

**\*Others:**
**Buzuruga HC (n=10):** *Morganella morganii* (n=3), *Enterobacter cloacae* complex (n=1), *Citrobacter koseri* (n=1), *Klebsiella oxytoca* (n=1), *Pseudomonas fluorescens* (n=1), *Pseudoglutamicibacter cumminsii* (n=1), *Staphylococcus epidermidis* (n=1), and *Staphylococcus haemolyticus* (n=1).
**Igoma HC (n=3):** *Enterobacter cloacae* complex (n=1), *Proteus mirabilis* (n=1), and *Streptococcus agalactiae* (n=1).

### Percentage resistance of bacterial pathogens causing CA-UTIs

*Escherichia coli* demonstrated the highest levels of antimicrobial resistance (≥50%) to multiple antibiotics, including ciprofloxacin, amoxicillin-clavulanic acid, trimethoprim-sulfamethoxazole, tetracycline, and ampicillin. About 20.3% of *E. coli* isolates showed positive phenotypes of extended-spectrum beta-lactamase (ESBL) production. Other members of the Enterobacterales group showed similarly high resistance rates to amoxicillin-clavulanic acid and ampicillin, with 12.0% detected with ESBL phenotypes. The non-fermentative Gram-negative bacteria, specifically *Acinetobacter* spp. and *Pseudomonas* spp., exhibited the highest resistance to amoxicillin-clavulanic acid, trimethoprim-sulfamethoxazole, ceftriaxone, and ampicillin (Table 4).

**Table 4:** Percentage resistance of Gram-negative bacteria causing community-acquired urinary tract infection at Buzuruga and Igoma Health Centers in Mwanza, Tanzania.

| Antibiotic agents tested | *E. coli* (N=64) |  | Other Enterobacterales (N=25) |  | Non-fermentative Gram-negative bacteria (N=27) |  |
| --- | --- | --- | --- | --- | --- | --- |
|  | % Resistance | % Sensitive | % Resistance | % Sensitive | % Resistance | % Sensitive |
| AMP | 58 (90.6) | 6 (9.4) | 25 (100) | 0 (0.0) | 5 (100) | 0 (0.0) |
| TET | 47 (73.4) | 17 (26.6) | 10 (40.0) | 15 (60.0) | 11 (47.8) | 12 (52.2) |
| SXT | 46 (73.0) | 17 (27.0) | 10 (40.0) | 15 (60.0) | 16 (59.3) | 11 (40.7) |
| CIP | 34 (53.1) | 30 (46.9) | 8 (32.0) | 17 (68.0) | 9 (33.3) | 18 (66.7) |
| NIT | 26 (40.6) | 38 (59.4) | 14 (56.0) | 11 (44.0) | 25 (92.6) | 2 (7.4) |
| AMC | 41 (64.1) | 23 (35.9) | 13 (52.0) | 12 (48.0) | 2 (50.0) | 2 (50.0) |
| CRO | 29 (45.3) | 35 (54.7) | 11 (44.0) | 14 (56.00) | 21 (80.8) | 5 (19.2) |
| GEN | 29 (45.3) | 35 (54.7) | 6 (25.0) | 18 (75.0) | 4 (15.4) | 22 (86.4) |
| MEM | 5 (7.9) | 58 (92.1) | 4 (16.0) | 21 (84.0) | 0 (0.0) | 26 (100) |
| ESBL strains | 13 (20.3) |  | 3 (12.0) |  | NA |  |

Key: AMP=ampicillin, TET=tetracycline, SXT=trimethoprim-sulfamethoxazole, CIP=ciprofloxacin, NIT=nitrofurantoin, AMC=amoxicillin-clavulanic acid, CRO=ceftriaxone, GEN=gentamicin, NA=not applicable, and MEM=meropenem.

On the other hand, *S. aureus* demonstrated markedly high resistance rates (≥ 50%) to ciprofloxacin, tetracycline, trimethoprim-sulfamethoxazole, ampicillin, and erythromycin. Moreover, 64.6% of *S. aureus* strains showed resistance towards cefoxitin, hence methicillin-resistant *S. aureus* (MRSA). Similarly, other Gram-negative bacteria exhibited peak resistance (≥ 50%) to ampicillin, tetracycline, trimethoprim-sulfamethoxazole, ciprofloxacin, and erythromycin (Table 3).

### Proportions of multidrug resistance and resistance patterns of Gram-negative bacteria causing CA-UTIs

Overall, the majority of Gram-negative bacteria (73.3%; 85/116) exhibited resistance to at least three classes of antimicrobials, with the number of classes ranging from three to six. By healthcare facility, the difference in proportions of MDR in Gram-negative bacteria isolated from patients attending at Buzuruga HC (70.5%; 43/61) and Igoma HC (76.4%; 42/55) was not statistically significant (p=0.475). *E. coli* showed the highest proportion of multidrug resistance (MDR), with 82.8% (53/64) of isolates classified as MDR, including 15.1% (n = 8) that were resistant to six antimicrobial classes. The common resistance patterns in *E. coli* were AMP-TET-SXT-CIP-NIT-AMC-CRO-CAZ-FEP-GEN-AK-TZP (n=3), AMP-TET-SXT-CIP-NIT-AMC-CRO-CAZ-FEP-GEN-TZP (n=2), and AMP-TET-SXT-CIP-AMC-CRO-CAZ-FEP-AK-TZP (n=2). Among other Enterobacterales, 44.0% (11/25) were MDR, with 27.3% (n = 3) also resistant to six classes. Similarly, 77.8% (22/27) of non-fermentative Gram-negative bacteria displayed MDR, with 9.1% (n = 2) resistant to six antimicrobial classes. The common resistance patterns in non-fermentative Gram-negative bacteria were SXT-NIT-CRO-CAZ-FEP (n=2) and TET-SXT-NIT-CRO-FEP-TZP (n=2) (Table 4). Detailed antimicrobial resistance patterns are provided in Supplementary File 1.

Among GPB, a substantial proportion of *S. aureus* isolates (83.3%; 40/48) were classified as MDR. Notably, 15.0% (6/40) of these MDR *S. aureus* strains exhibited resistance to as many as seven antibiotics. MDR phenotypes were significantly more prevalent among *S. aureus* isolates from patients at Igoma Health Centre compared to those from Buzuruga HC (95.6% vs. 72.0%; p = 0.019). The most frequently observed resistance patterns in *S. aureus* included CD-ER-AMP-SXT-TET-CIP-FOX (n=3), ER-AMP-SXT-TET-CIP-FOX (n=3), and ER-AMP-SXT-TET-NIT-CIP-FOX-GEN (n=3). For other GPB, all isolates (100%; 29/29) were MDR. The most common resistance profiles among these included CD-ER-AMP-SXT-TET-CIP (n=2), CD-ER-NIT-CIP (n=2), and ER-AMP-SXT-TET-NIT-CIP (n=2). Moreover, 24.1% (7/29) of these MDR GPB strains demonstrated resistance to six antimicrobial agents (Table 5). Detailed antimicrobial resistance patterns are shown on Supplementary file 2.

**Table 5:** Percentage resistance of Gram-positive bacteria causing community-acquired urinary tract infection at Buzuruga and Igoma Health Centers in Mwanza, Tanzania.

| Antibiotic agents tested | *S. aureus* (N=48) |  | Other Gram-positive bacteria (N=29) |  |
| --- | --- | --- | --- | --- |
|  | % Resistance | % Sensitive | % Resistance | % Sensitive |
| ER | 43 (89.6) | 5 (10.4) | 28 (96.6) | 1 (3.4) |
| CIP | 24 (50.0) | 24 (50.0) | 25 (86.2) | 4 (13.8) |
| SXT | 31 (64.6) | 17 (35.4) | 24 (82.8) | 5 (17.2) |
| TET | 28 (58.4) | 20 (41.6) | 22 (75.9) | 7 (24.1) |
| PEN | 38 (79.2) | 10 (20.8) | 15 (51.7) | 14 (48.3) |
| CD | 29 (31.3) | 33 (68.7) | 14 (48.3) | 15 (51.7) |
| GEN | 11 (22.9) | 37 (77.1) | 11 (37.9) | 18 (62.1) |
| NIT | 12 (25.0) | 36 (75.0) | 11 (37.9) | 18 (62.1) |
| FOX | 31 (64.6) | 17 (35.4) | NT | NT |

Key: ER=erythromycin; CIP=ciprofloxacin; SXT=trimethoprim-sulfamethoxazole; TET=tetracycline; PEN=penicillin; CD=clindamycin; GEN=gentamicin; NIT-nitrofurantoin; and FOX=cefoxitin.

**Table 6:** Proportions of multidrug resistance in Gram-negative bacteria causing community-acquired urinary tract infection at Buzuruga and Igoma Health Centers in Mwanza, Tanzania.

| Variable | Category | *E. coli*<br>(N=64) | Other Enterobacterales<br>(N=25) | Non-fermentative<br>GNB (N=27) |
| --- | --- | --- | --- | --- |
|  |  | n(%) | n(%) | n(%) |
| Multidrug-resistant<br>(MDR) strain | Yes | 53 (82.8) | 11 (44.0) | 21 (77.8) |
|  | No | 11 (17.2) | 14 (56.0) | 6 (22.2) |
| Number of antibiotic<br>classes the bacteria<br>resist | 3 | 10 (18.9) | 2 (18.2) | 10 (45.4) |
|  | 4 | 13 (24.5) | 4 (36.4) | 6 (27.3) |
|  | 5 | 22 (41.5) | 2 (18.2) | 3 (13.6) |
|  | 6 | 8 (15.1) | 3 (27.3) | 2 (9.1) |
| Health centre of<br>isolation of MDR strains | Buzuruga HC<br>(N=43) | 28 (65.1) | 9 (20.9) | 6 (13.9) |
|  | Igoma HC (N=42) | 25 (59.5) | 12 (28.6) | 5 (11.9) |

**Table 7:**
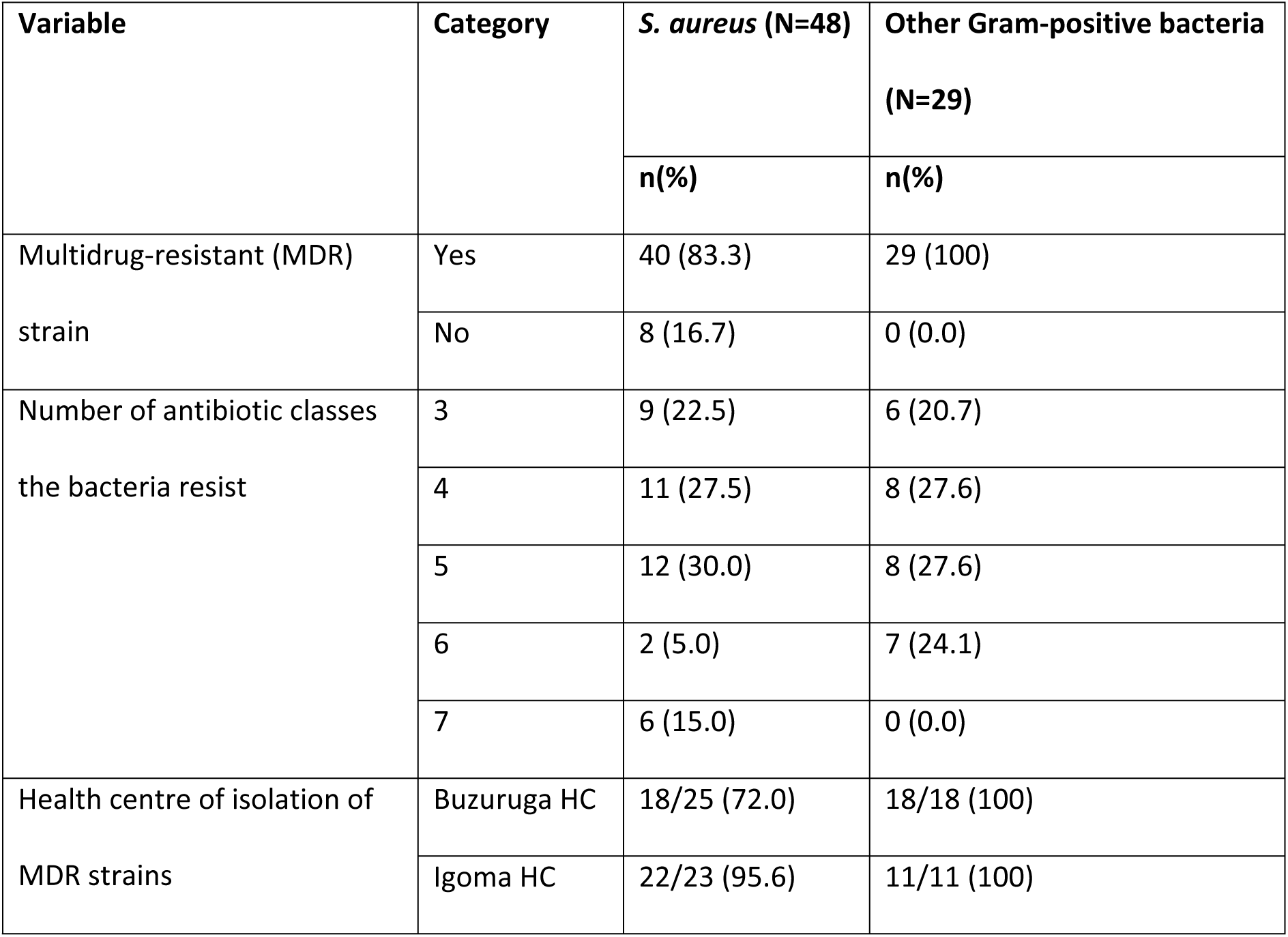
Proportions of multidrug resistance in Gram-positive bacteria causing community-acquired urinary tract infection at Buzuruga and Igoma Health Centers in Mwanza, Tanzania.

## Discussion

Urinary tract infection is currently a public health concern especially when acquired from community settings due to its high prevalence, increasingly drug-resistant, costly to treat, and associations with serious complications if not managed effectively. The disease is reported to be among the leading complain among patients attending outpatient’s clinic[21]. It has gained more attention recently due to antimicrobial resistance associated with pathogens causing UTI that limit treatment option especially in lower- and middle-income settings. This study was conducted among community members attending the lower health facility with clinical signs and symptoms of UTI.

The prevalence of community-acquired, microbiologically confirmed UTI in the present study was 22%, which falls within the previously reported national range of 11–23% from diverse populations in Tanzania [5–7]. However, this prevalence is slightly lower than the 27% reported by Silago et al. in the same setting three years earlier [9]. This discrepancy may be explained by differences in study populations; the earlier study included pregnant women a group with a physiologically higher susceptibility to UTI while the current study excluded them.

From a public health perspective, a 22% prevalence indicates that UTIs remain a substantial community health burden, contributing to frequent outpatient visits and antimicrobial prescriptions. This underscores the need for strengthened community-level infection prevention strategies, including improved personal hygiene practices, safe water access, and health education aimed at early care seeking and avoidance of self-medication[22]. Importantly, most participants reported prior antibiotic use, a finding with significant implications. Such widespread community antibiotic exposure often unregulated can suppress bacterial growth and potentially lead to underestimation of true UTI prevalence while simultaneously exerting selection pressure that drives antimicrobial resistance (AMR).

In the current study, just like other studies, Gram-negative bacteria were predominant pathogens detected causing community acquired UTI. The predominance of Gram-negative bacteria is partly contributed by the fact that these bacteria are normal microbiota of the gastrointestinal tract and the close proximity of the urinary and gastrointestinal tract make it easy for the organism to ascend and cause urinary tract infection.

The predominance of Gram-negative bacteria, especially *E. coli*, is consistent with global and regional trends in community-acquired urinary tract infections (CA-UTIs), reflecting its well-established role as the primary uropathogen [9]. The notable presence of *S. aureus*, a Gram-positive organism, suggests potential shifts or local variations in CA-UTI causes, warranting further exploration. Differences in less common isolates between the two health centres may be linked to local antimicrobial use, patient demographics, or environmental factors influencing bacterial populations. The exclusive detection of *C. freundii* at one site highlights potential micro-epidemiological differences, which could inform more targeted infection control and surveillance efforts.

The high resistance observed, particularly among *E. coli* and non-fermentative Gram-negative bacteria, emphasizes the increasing challenge of treating CA-UTIs with commonly prescribed antibiotics [23]. These findings underscore the urgent need for ongoing surveillance, rational antimicrobial use, and updated treatment guidelines tailored to local resistance patterns to manage infections effectively and limit the spread of multidrug-resistant pathogens. The widespread multidrug resistance (MDR) observed in Gram-negative bacteria, with *E. coli* showing the highest levels, is particularly concerning, given its dominance as a uropathogens. Resistance to multiple antibiotic classes, including critical agents like piperacillin-tazobactam and amikacin, limits treatment options and reinforces the critical need for antimicrobial stewardship to address this growing threat.

The high MDR prevalence among *S. aureus* (83.3%) and other GPB (100%) in this study is consistent with findings by Silago et al. in Mwanza and Dar es Salaam, where similar resistance patterns to ciprofloxacin, tetracycline, and trimethoprim-sulfamethoxazole were observed [9]. Notably, the resistance profiles align with post-COVID-19 trends reported by the same group, showing persistent and widespread MDR uropathogens in Tanzania [24]. The significantly higher MDR burden at Igoma HC may reflect differences in local antimicrobial practices. These findings underscore the ongoing threat of resistant GPB pathogens and the urgent need for context-specific antimicrobial stewardship in outpatient settings. The resistance patterns observed in this study may therefore reflect ongoing community misuse or overuse of antibiotics previously reported [25, 26], reinforcing the urgency of antimicrobial stewardship interventions, regulation of non-prescription antibiotic sales, and public education on the risks of inappropriate antibiotic use.

## Conclusion and Recommendation

### Conclusion

This study revealed a significant burden of community-acquired urinary tract infections (CA-UTIs) among adult outpatients in Mwanza, Tanzania, with an overall prevalence of 22%. *Escherichia coli* and *Staphylococcus aureus* were the most frequently isolated pathogens, and alarmingly high levels of antimicrobial resistance particularly multidrug resistance in *E. coli* were observed. The late presentation of patients, limited recognition of classical UTI symptoms, and high recurrence rates further complicate clinical management.

### Recommendations

Routine microbiological surveillance should be established in outpatient settings to monitor the evolving antimicrobial resistance patterns and inform empirical treatment guidelines. Additionally, public health education campaigns are needed to raise awareness about early UTI symptoms, discourage self-medication, and encourage prompt medical consultation. Enhancing diagnostic capacity at primary healthcare facilities, including the availability of urine culture and sensitivity testing, will improve timely diagnosis and targeted treatment. Finally, national health authorities should consider updating and disseminating evidence-based treatment protocols for CA-UTIs that reflect local resistance trends and support implementation across all levels of care.

## Data Availability

All data are included in the manuscript

